# Residency in the Era of Climate Change: A Multi-Institutional Survey of Medical Student Perceptions and Match Preferences

**DOI:** 10.64898/2026.01.25.26344334

**Authors:** Caroline J. Walsh, Emma R. Shelby, Cameron Quick, Kate Hanneman, Robert K. Ryu, Reed A. Omary

## Abstract

**Background:** Climate change is an escalating health crisis, yet its influence on medical students’ career decisions remains underexplored.

**Objective:** This study uses the “Six Americas” framework to assess students’ climate views and the impact on residency and career decisions.

**Methods:** A survey was distributed to students at four North American medical schools from October 2024 to January 2025. The instrument assessed demographic information, climate change views utilizing the Six Americas Super Short Survey, and the impact of climate change on residency and career decisions. Respondents were classified as Alarmed, Concerned, Cautious, Disengaged, Doubtful, or Dismissive using the Six Americas framework. Associations between climate attitudes, training year, and intended specialty were analyzed using Kruskal-Wallis tests.

**Results:** A total of 105 medical students completed the survey. 93% agreed climate change is a major health threat, 66% valued residency program sustainability efforts, and 41% indicated commitment to sustainability influences their rank list. Most students were Alarmed (62%), Concerned (19%), or Cautious (12%). Climate attitudes did not vary by specialty interest. Training year was associated with Six Americas classification (p < 0.05), MS2s had the most Alarmed respondents.

**Conclusions:** Medical students overwhelmingly view climate change as a serious health threat and expect residency programs to demonstrate commitment to sustainability. Nearly half consider environmental values when ranking programs. These findings suggest climate-conscious training environments may gain a competitive recruitment advantage. Given the high proportion of “Alarmed” students, embedding climate-resilient education and sustainable practices into residency programs may align with future physician priorities.

## Introduction

The impacts of climate change on health are increasingly recognized as a critical concern, including among medical students. Prior studies demonstrate medical students view climate change as a health crisis and advocate that planetary health education be more strongly integrated ^1–5^. Globally, students express a strong sense of responsibility, but gaps persist in climate-related knowledge and training ^4–7^.

While graduate medical programs have begun incorporating climate topics into curricula, it remains unclear whether students consider environmental values when selecting residency programs ^8,9^. As applicants increasingly weigh factors like reproductive rights and institutional culture, it is plausible that climate-conscious values may also shape residency decisions ^10–14^.

A 2020 Princeton Review survey reported that 56% of prospective college students considered environmental commitment an important factor in school selection ^15^. This trend suggests that many of today’s medical students—former undergraduates—may similarly prioritize sustainability when evaluating future training.

The objective of our study was to examine medical students’ attitudes on climate change and to understand how climate change affects medical student preferences for residency program and specialty selection. We tested the hypothesis that medical students consider climate change an important factor in residency program selection, thereby influencing career decisions.

## Methods

### Participant Characteristics

This study employed a cross-sectional survey design to assess medical student attitudes toward climate change and its influence on residency program selection.

### Survey Characteristics

Due to the lack of existing validated instruments addressing medical student climate attitudes in the context of residency selection, we developed our survey based on a review of relevant literature ^12,13,16–20^. The initial survey draft was created by the research team and reviewed by three faculty mentors with expertise in medical education and sustainability. Revisions were made and the instrument was piloted among five medical students to ensure clarity and readability. We intentionally developed a short survey to increase response rate and optimize data quality ^21^.

The final survey included a total of ten questions (Supplemental Table 1). Surveys were de-identified; no sensitive demographic questions were included. The first two questions assessed medical school year and intended residency specialty. Prior to data analysis, intended specialties were consolidated into broader categories due to a low number of respondents in many individual specialties. The next four questions assessed student beliefs, attitudes, and behaviors regarding climate change in medical education and residency selection using a five-point Likert scale (strongly disagree to strongly agree) to standardize responses and facilitate quantitative analysis ^22^.

The final four questions assessed respondent’s attitudes towards climate change using a standardized tool, the “Six Americas Super Short Survey” (SASSY) developed by the Yale Program on Climate Change Communication^23^. The data were coded and entered in the SASSY Group Scoring Tool which grouped each respondent into one of the subgroups: “Alarmed,” “Concerned,” “Cautious,” “Disengaged,” “Doubtful,” and “Dismissive.” The Six Americas segmentation was originally based on 36 items and Latent Class Analysis. The short version of the SASSY uses just four items and classifies respondents using a multinomial logistic regression model trained on prior national survey data. In this model, the dependent variable is the six subgroups while the four items serve as predictors. The model estimates odds ratios for each response option which then are used to calculate the probability of membership in each segment; respondents are assigned to the category with the highest predicted probability. Compared with the original 36-item latent class model, the short survey correctly classified between 70-87% of respondents into the same segment across multiple validation datasets ^23^. Applying this framework allows a nuanced analysis of where medical students fall on the spectrum of climate concern spectrum.

According to Yale’s Program on Climate Change Communication, both the “Alarmed” and “Concerned” groups agree that human-caused climate change is occurring and a serious threat while supporting climate policies. In the present survey, survey respondents who fell into either group were identified as “Climate Committed.” Conversely, the “Doubtful,” and “Dismissive” group do not believe climate change is happening and caused by humans. Thus, we labelled these two as “Climate Non-Committed.” No sub-groups such as these have been proposed in the past. The “Cautious” and “Disengaged” groups were not included in either sub-group as they do not have formed opinions on climate change; they are either skeptical that climate change exists and is human caused (cautious) or know little about climate change (disengaged).

The survey was distributed via email to medical students at four academic institutions—three located in the United States and one in Canada. Survey links were distributed in October 2024 and data were collected through January 2025. Email scripts were standardized across schools and approved by the student data advisory team (or equivalent review board) at the discretion of each institution.

### Statistical Analysis

Descriptive statistical analysis was performed in Microsoft Excel. To assess differences in climate change attitude and its impact on residency program selection across specialty sub-group interests and medical school years, we conducted Kruskal-Wallis One-Way ANOVA tests followed by Dwass-Steel-Critchlow-Fligner (DSCF) pairwise comparisons for post-hoc analysis. The same methodology was used to evaluate for associations between specialty sub-group interest and medical school year and Six Americas group. All analyses were performed on the free and open statistical platform, Jamovi ^24^. Statistical significance was set at p < 0.05. The DSCF test inherently adjusts for multiple comparisons in pairwise testing following a significant Kruskal-Wallis test. We utilized a chi-square test of independence to assess whether the proportion of students who felt alarmed differed by specialty and medical school years. Statistical significance was set at p < 0.05.

### Institutional Review Board

The University of Colorado Multiple Institutional Review Board deemed the study exempt from review (COMIRB #24-1428).

## Results

A total of 105 medical students completed the survey. Seven (6.7%) were from a single Canadian institution, while the remaining 98 (93.3%) were from three institutions in the United States. Respondent characteristics are summarized in **Table 1**. The distribution of intended specialties among respondents is illustrated in **Figure 1**.

**Figure 1.**
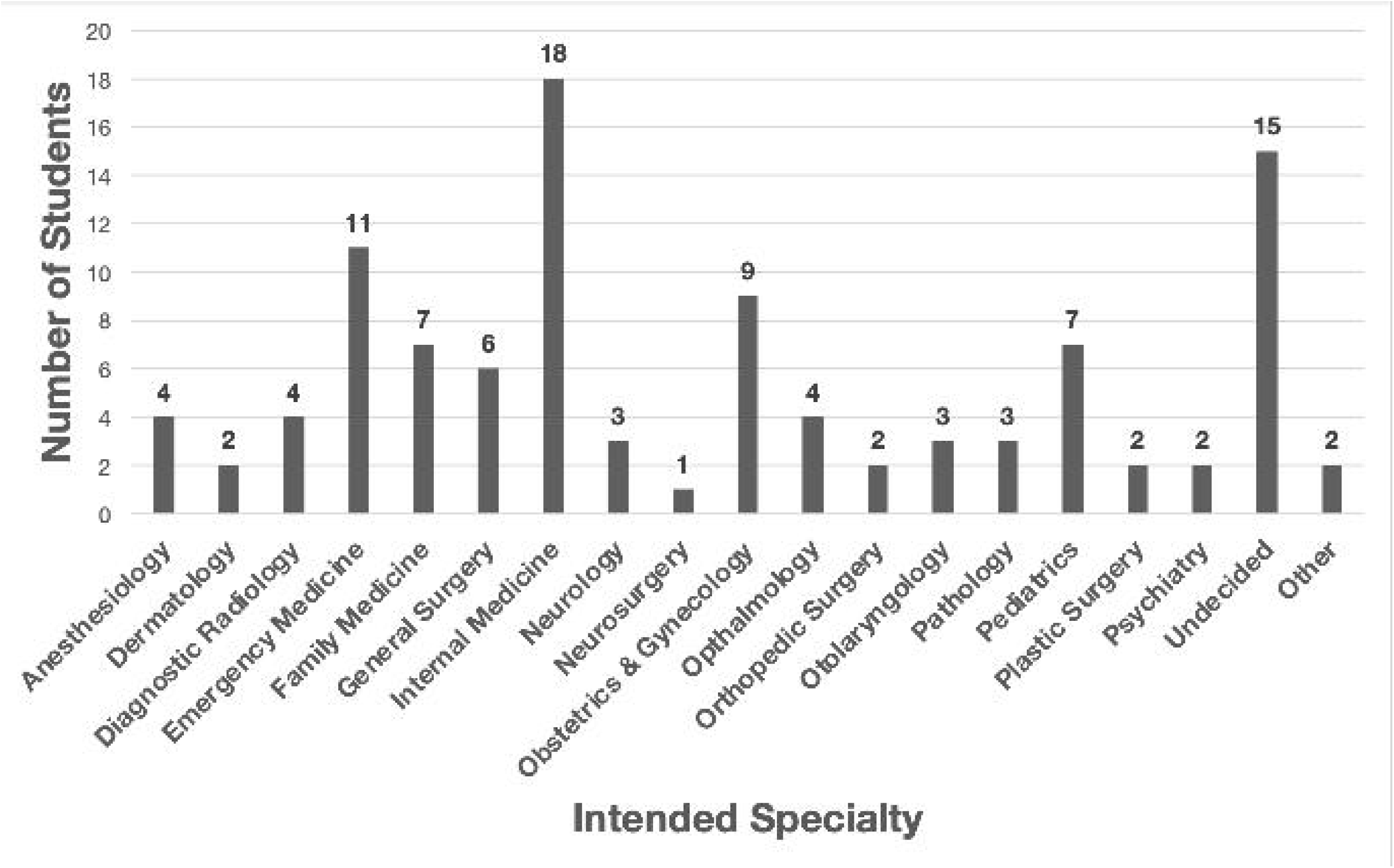
Distribution of intended specialty selection among survey respondents. Specialties with no respondents were not included in the figure (this includes interventional radiology, medical genetics and genomics, physical medicine and rehabilitation, preventative medicine, radiation oncology, thoracic surgery, and urology).

**Table 1.** Survey respondent characteristics. Primary care included: pediatrics, FM, IM, OB/Gyn. Surgical included: otolaryngology, orthopedic surgery, neurosurgery, ophthalmology, and general surgery. Medical specialties included: neurology, diagnostic radiology, dermatology, anesthesiology, emergency medicine, psychiatry, and pathology.

| Survey Categories | Number of Students (n = 105) |
| --- | --- |
| **Medical School** |  |
| US School 1 | 50 |
| US School 2 | 28 |
| US School 3 | 20 |
| Canadian School | 7 |
| **Year in Medical School** |  |
| 1 | 35 |
| 2 | 14 |
| 3 | 28 |
| 4 | 28 |
| **Intended Specialty Category** |  |
| Primary Care | 41 |
| Surgical | 18 |
| Medical Specialists | 29 |
| Undecided | 17 |

A total of 93% of respondents agreed or strongly agreed with the statement “climate change is an important threat to human health,” while only 3% disagreed or strongly disagreed (**Figure 2**). 42% of medical students agreed or strongly agreed that they will consider the importance of climate change and its impact on their career and patients when choosing a specialty (**Figure 2**). Two-thirds (66%) of respondents agreed or strongly agreed that it is important for their residency program to acknowledge climate change and demonstrate a commitment to sustainability, compared to 15% who disagreed or strongly disagreed. Finally, 41% of medical students reported that a program’s dedication to climate change mitigation will impact their residency program preference list (**Figure 2**).

**Figure 2.**
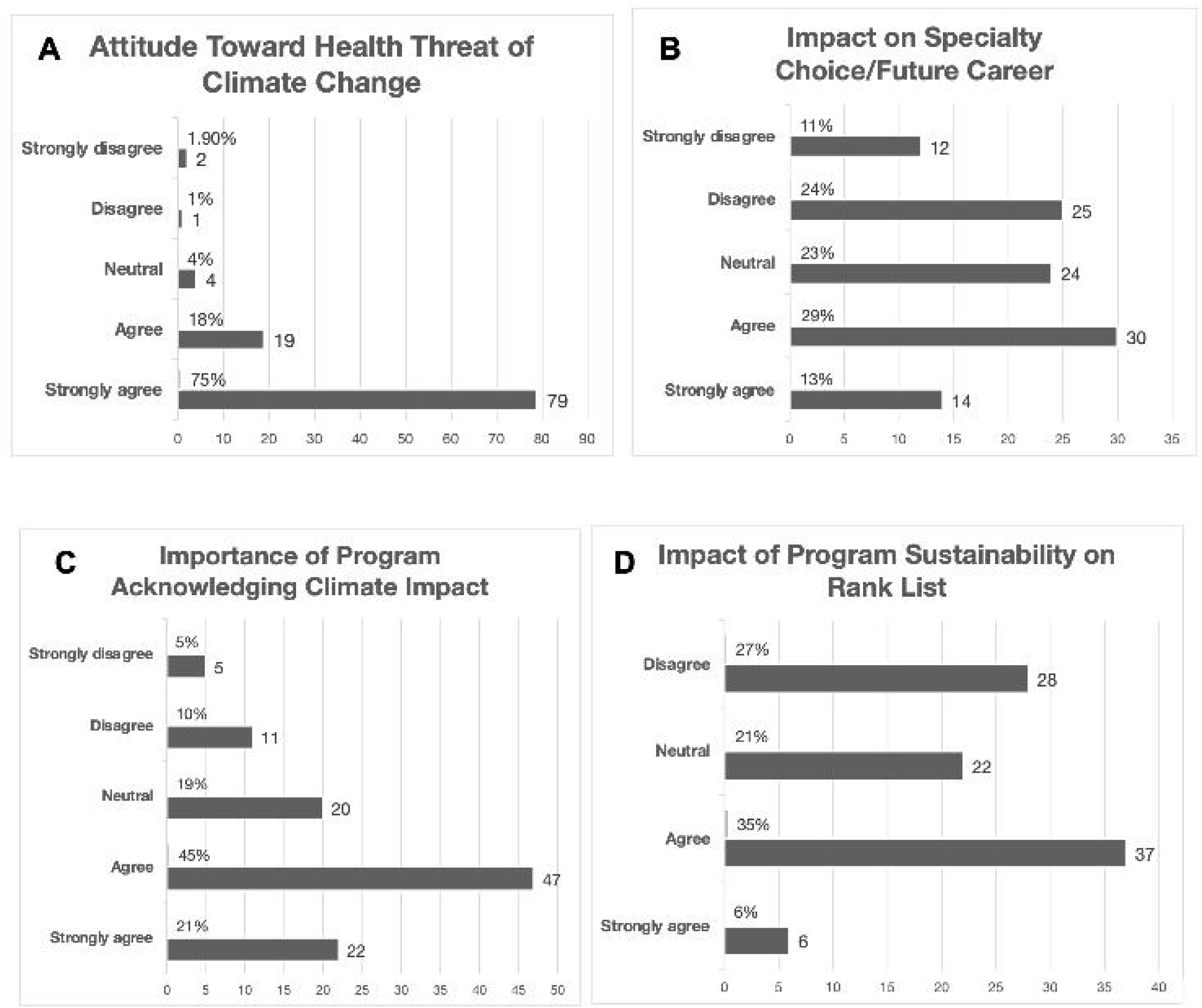
Percent of medical students that strongly agreed, agreed, felt neutral, disagreed, or strongly disagreed with each survey question (a-d below) assessing student attitude toward climate change (*number of respondents written to the right of the bar and percentage out of 105 written above respective bar*). a) “Climate change is an important threat to human health”; b) “In choosing my medical specialty, I consider the relative importance of climate change, and the potential impact it will have on my career and patients”; c) “It is important to me that my residency program acknowledges the health impact of climate change and demonstrates commitment to sustainability”; and d) “The level of dedication to sustainability and climate change mitigation will impact my preference list of residency programs.”

Based on survey responses to the four questions from the Six Americas Super Short Survey (SASSY!), 62% of medical students were classified as “Alarmed” in the Global Warming’s Six Americas Framework. Overall, 81% of medical students were categorized into either the “Alarmed” or “Concerned” group and thus considered “Climate Committed.” Only 5% were categorized as either “Doubtful,” or “Dismissive” and thus considered “Climate Non-Committed” (**Table 2 and Figure 3**).

**Figure 3.**
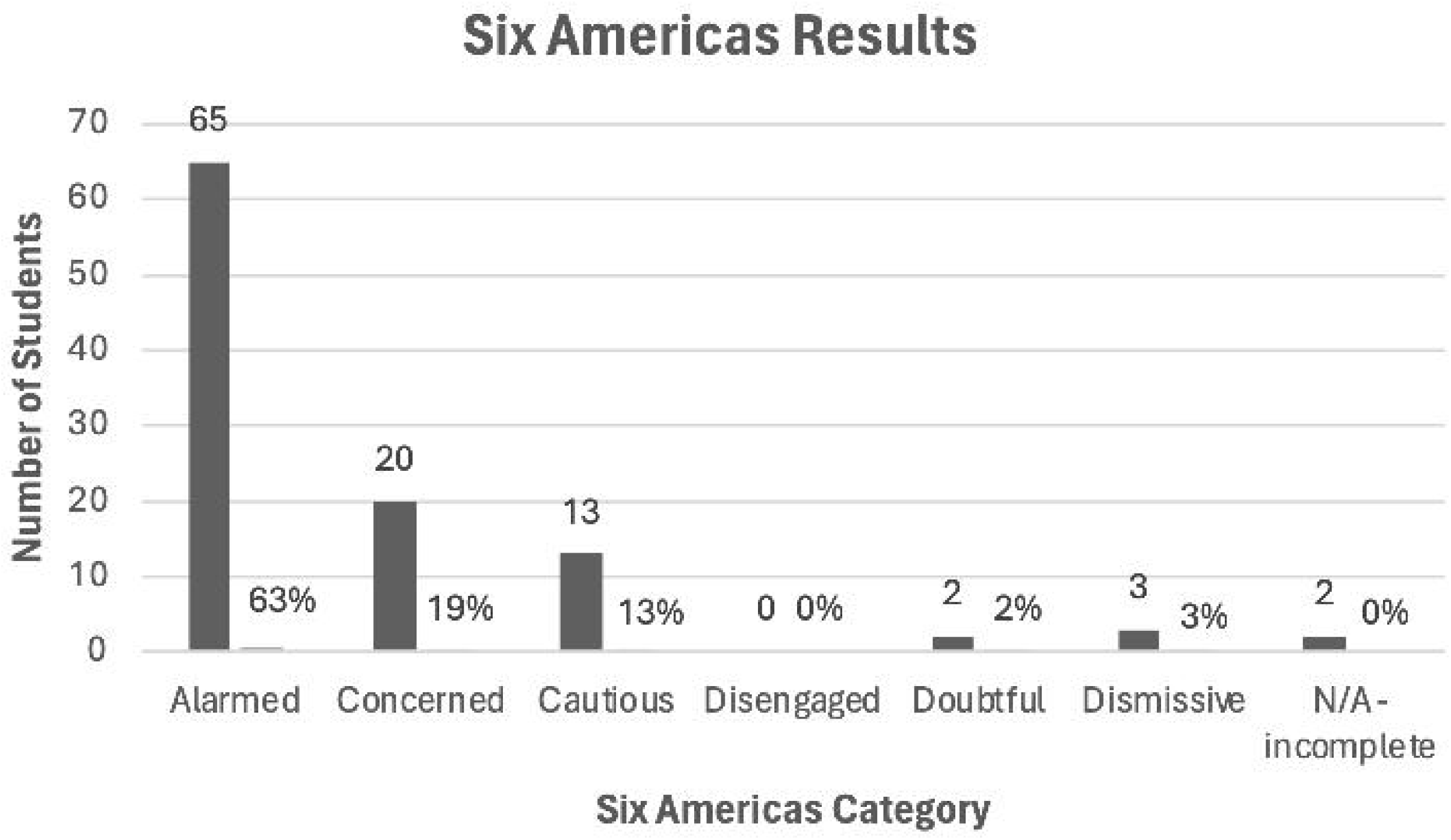
Distribution of Global Warming’s Six America Results Among Medical Student Respondents

**Table 2.** Number of Medical Students in Each Group of the Global Warming’s Six America.

|  | Alarmed | Concerned | Cautious | Disengaged | Doubtful | Dismissive | N/A |
| --- | --- | --- | --- | --- | --- | --- | --- |
| **TOTAL (105)** | 65 (62%) | 20 (19%) | 13 (12%) | 0 (0%) | 2 (2%) | 3 (3%) | 2 (2%) |
| **Med School Year** |  |  |  |  |  |  |  |
| MS1 (35) | 16 (46%) | 8 (23%) | 6 (17%) | 0 (0%) | 2 (6%) | 2 (6%) | 1 (3%) |
| MS2 (14) | 12 (86%) | 1 (7%) | 1 (7%) | 0 (0%) | 0 (0%) | 0 (0%) | 0 (0%) |
| MS3 (28) | 20 (71%) | 4 (14%) | 4 (14%) | 0 (0%) | 0 (0%) | 0 (0%) | 0 (0%) |
| MS4 (28) | 18 (64%) | 7 (25%) | 2 (7%) | 0 (0%) | 0 (0%) | 1 (4%) | 0 (0%) |
| **Specialty** |  |  |  |  |  |  |  |
| Primary Care (41) | 27 (66%) | 7 (17%) | 5 (12%) | 0 (0%) | 1 (2%) | 0 (%) | 1 (2%) |
| Surgical (18) | 11 (61%) | 4 (22%) | 1 (6%) | 0 (0%) | 0 (0%) | 2 (11%) | 0 (0%) |
| Med Specialties (29) | 18 (62%) | 6 (21%) | 5 (17%) | 0 (0%) | 0 (0%) | 0 (0%) | 0 (0%) |
| Undecided (17) | 10 (59%) | 3 (18%) | 2 (12%) | 0 (0%) | 1 (6%) | 1 (6%) | 0 (0%) |

*Note: Total number of students in each category are listed in parenthesis in the first column. Percentage of students within each category (MS1, MS2, MS3, etc.) that fall within each subgroup is listed in parenthesis in columns 2-8.*

There was no significant association between intended medical specialty and attitude toward health threat of climate change (χ²(3)= 1.69, p= 0.64), impact on specialty choice (χ²(3)=7.31, p= 0.063), importance of program acknowledging climate impact (χ²(3)=6.2, p= 0.10), or impact of program sustainability on rank list (χ²(3)= 5.74, p= 0.12). Similarly, there was no significant differences in the Six Americas classifications across specialty interest groups (χ²(3)= 0.414, p= 0.94). Post-hoc pairwise comparisons using the DSCF test confirmed no statistically significant differences between any specialty subgroup pairs. The distribution of “Alarmed” students did not differ significantly by subspecialty interest group (χ²(3) =0.307, p = 0.96).

There were no significant associations between medical school year and attitude towards climate change or its influence on residency program selection. Conversely, there was a difference in Six Americas group distribution across the four medical school years (χ²(3) =9.6, p= 0.022). However, post-hoc pairwise comparisons revealed no differences between any individual years (all p> 0.05). The distribution of “Alarmed” students differed significantly by medical school year (χ²(3) = 8.44, p = 0.038); MS2s had the greatest number of Alarmed students (86%) followed by MS3s (71%), MS4s (64%), and MS1s (46%) (**Table 2**).

## Discussion

In this multi-institutional survey, the vast majority of medical students agreed climate change poses a significant threat to human health. Most reported the importance of residency programs to acknowledge climate impacts and highlight sustainability efforts. Many students felt climate-related factors would influence their residency program rankings or career decisions. Most respondents were “Climate Committed.”

### Medical Student Attitudes Toward Climate Change

Our results reinforce prior studies showing that medical trainees and US youth strongly perceive climate change as a health threat. For example, Lewandowski et al. surveyed 15,793 U.S youth and found widespread climate-related distress with 85% at least moderately worried and 58% very or extremely worried with 40% reporting it harms mental health ^25^. Similarly, among German medical students, Bugaj et al. found the majority of students expected health impacts of climate change (76%) and felt individual responsibility (75%)^7^. Heydari et al. and Ryan et al. revealed similarly high concern for climate change among Iranian medical students and US medical, nursing, and PA students, with a large interest to increase education^5,26^. These findings reflect a global generation of youth and health professional trainees deeply engaged with climate disruption and motivated to act.

Prior research has centered on medical trainee knowledge of climate health impacts or desire for planetary health curriculum. We explored a new direction, assessing potential differences in attitude by year in training and intended specialty. While no subgroup differences reached significance, the consistently high concern reinforces the trend in medical students that climate change is urgent.

### Application of Six Americas to Medical Students

This study introduces a novel application the Six Americas framework—a validated, widely used tool for assessing climate change attitudes across diverse populations—by examining the tool in medical students. This model, commonly used in public opinion research, offers a structured, scalable lens to inform future studies across specialties, training levels, and regions.

In the present study, most respondents were categorized as either “Alarmed” (62%) or “Concerned” (19%) and therefore classified as “Climate Committed”, which is substantially greater than the general public (26%, and 28% respectively) ^27^. Only 5% were classified as “Climate Non-Committed.” While the United States is the largest cumulative emitter of carbon, it contains some of the largest populations of “Climate Non-Committed” individuals (25%) ^28^. These findings and the increased prevalence of “Climate Committed” medical students suggest medical trainees may have stronger alignment with climate science than the public, paralleling trends of increasing alarm among youth cohorts globally ^25,29^.

Over the past decade, the “Cautious” group has decreased in size the most, representing just 18% in 2024 ^29^. In our study, they represent only 12% of students. The documented shift from the “Cautious” to “Concerned” or “Alarmed” groups is likely due to increased education, increased scientific research linking climate change to chronic diseases, and growing exposure to extreme weather ^30–32^. Globally, the “Alarmed” group has grown the most. More respondents are gaining concern for climate change than the opposite, consistent with our findings of 62% of “Alarmed” students ^29,33^. This raises the question of whether concern is specific to younger generations in medicine or persists across career stages. Extending this standardized questionnaire to health professionals across career stages could illuminate how climate attitudes evolve and where educational interventions might have the greatest impact.

### Impact of Climate Change on Residency Selection and Career

A key finding from this study is the potential influence of climate change on medical students’ career choices. Nearly half of students report climate-related factors will influence their selection of a residency program and most report that it is important their residency program acknowledges the health impact of climate change and commits to sustainable practices. Nearly half of students consider the importance of climate change and the impact it will have on their career and patients.

These findings suggest that environmental values inform professional decisions, whether through choosing programs with sustainable practices and planetary health curricula, seeking locations less vulnerable to climate impacts, prioritizing specialties with relevance to environmental health (e.g., emergency medicine, psychiatry, or pediatrics), or finding environmental stewardship as integral to professional identity ^1–4,7,8,26,34,35^. For example, a large international survey by Omrani et al. found that medical students across 112 countries strongly advocate to integrate climate change and planetary health topics into medical curricula, with many students actively seeking programs that address these issues and desiring meaningful involvement in curriculum transformation ^36^.

Our data suggest medical students look to residency programs to teach about climate change and health, train climate-competent physicians who mitigate healthcare’s environmental impact, and foster sustainability initiatives, findings supported by previous research ^9,37^. Beyond training, healthcare professionals seek to address the impact of climate change on health and desire increased education ^38^. We postulate residency programs that prioritize climate-conscious curriculum and sustainable practices will have a recruiting advantage among students who value sustainability.

### Study limitations

This study has several important limitations. First, data were collected from medical students at only four institutions, mostly within the US. While not all US regions were included, respondents likely represent a diverse sample, as many students attend institutions outside their home region. Second, the study may be subject to selection bias as respondents may disproportionately represent individuals with preexisting interest or engagement in the topic of climate change. This may have contributed to the low proportion of “Dismissive” and “Disengaged” students. The small sample size likely also led to underpowered comparisons, explaining why no significant findings were observed across sub-specialty groups. Finally, in keeping our survey brief, we did not assess demographic factors such as race, age, and gender^21^. Evidence suggests women and youth are more invested in planetary health, future research could explore demographics more deeply^25,39,40^.

## Conclusions

This study suggests medical students are highly concerned about climate change while offering new insight into how these concerns impact residency selection. Medical students are more alarmed by and committed to addressing climate change than the public, with expectation residency programs commit to sustainability. Many report this commitment will impact their residency program rank list. Thus, we recommend residency programs prioritize climate-conscious policies, invest in a climate-conscious culture and education, and engage in mitigation efforts. Future studies might assess which sustainability initiatives student applicants value most and measure how residency programs respond to this growing demand. As the health impacts of climate change intensify, medical institutions that align with the values of incoming trainees will remain competitive, relevant, and responsive to global health needs. Supporting climate-conscious education and infrastructure is therefore both a public health imperative and a wise recruitment strategy.

## Acknowledgements

None.

## Disclaimer

None.

## Ethical approval

IRB exemption was obtained.

## Data Availability

Data are available from the corresponding author upon reasonable request and with permission from University of Colorado’s IRB, in accordance with institutional policies.

## References

1. Hampshire K, Ndovu A, Bhambhvani H, Iverson N. Perspectives on climate change in medical school curricula—a survey of US medical students. The Journal of Climate Change and Health. 2021;4:100033.

2. Brouillette M. Medical Schools Are Updating Their Curricula as Climate Change Becomes Impossible to Ignore. JAMA. Sep 10 2024;332(10):775–776. doi:10.1001/jama.2024.13506

3. Rybol L, Nieder J, Amelung D, et al. Integrating climate change and health topics into the medical curriculum - a quantitative needs assessment of medical students at Heidelberg University in Germany. GMS J Med Educ. 2023;40(3):Doc36. doi:10.3205/zma001618

4. Liao W, Yang L, Zhong S, et al. Preparing the next generation of health professionals to tackle climate change: Are China’s medical students ready? Environ Res. Jan 2019;168:270–277. doi:10.1016/j.envres.2018.10.006

5. Ryan EC, Dubrow R, Sherman JD. Medical, nursing, and physician assistant student knowledge and attitudes toward climate change, pollution, and resource conservation in health care. BMC Med Educ. Jun 23 2020;20(1):200. doi:10.1186/s12909-020-02099-0

6. Pandve HT, Raut A. Assessment of awareness regarding climate change and its health hazards among the medical students. Indian J Occup Environ Med. Jan 2011;15(1):42–5. doi:10.4103/0019-5278.82999

7. Bugaj TJ, Heilborn M, Terhoeven V, et al. What do Final Year Medical Students in Germany know and think about Climate Change? - The ClimAttitude Study. Med Educ Online. Dec 2021;26(1):1917037. doi:10.1080/10872981.2021.1917037

8. Peterson TD, Domingo A, Stadler D, et al. An Interprofessional Approach to Prepare Medical Residents and Fellows to Address Climate- and Environment-Related Health Risks. J Grad Med Educ. Dec 2024;16(6 Suppl):5–10. doi:10.4300/JGME-D-24-00109.1

9. Moon C, Braganza S, Bathory E. Incorporating Climate Change Education Into Residency: A Focus on Community Risks and Resources. J Grad Med Educ. Dec 2024;16(6 Suppl):86–91. doi:10.4300/JGME-D-24-00061.1

10. Fereydooni A, Ramirez JL, Morrow KL, Chandra V, Coleman DM, Lee JT. Factors influencing medical student choices in the integrated vascular surgery match: Implications for future post-pandemic residency matches. J Vasc Surg. Oct 2021;74(4):1354–1361.e4. doi:10.1016/j.jvs.2021.05.014

11. Ladha FA, Pettinato AM, Perrin AE. Medical student residency preferences and motivational factors: a longitudinal, single-institution perspective. BMC Med Educ. Mar 17 2022;22(1):187. doi:10.1186/s12909-022-03244-7

12. Hulsman L, Bradley PK, Caldwell A, Christman M, Rusk D, Shanks A. Impact of the Dobbs v Jackson Women’s Health Organization decision on retention of Indiana medical students for residency. Am J Obstet Gynecol MFM. Nov 2023;5(11):101164. doi:10.1016/j.ajogmf.2023.101164

13. Mermin-Bunnell K, Traub AM, Wang K, Aaron B, King LP, Kawwass J. Abortion restrictions and medical residency applications. J Med Ethics. Jan 23 2025;51(2):79–86. doi:10.1136/jme-2023-109190

14. Hammoud MM, Morgan HK, George K, et al. Trends in Obstetrics and Gynecology Residency Applications in the Year After Abortion Access Changes. JAMA Netw Open. Feb 05 2024;7(2):e2355017. doi:10.1001/jamanetworkopen.2023.55017

15. Krier J. College Hopes & Worries Survey. The Princeton Review; 2025. p. 1–10.

16. Lee KE, Lim F, Silver ER, Faye AS, Hur C. Impact of COVID-19 on residency choice: A survey of New York City medical students. PLoS One. 2021;16(10):e0258088. doi:10.1371/journal.pone.0258088

17. Benedict MD, Hespe GE, Kumar NG, Xi AS, Myers PL, Sears ED. The Impact of Social Media on Applicants’ Perceptions of Plastic Surgery Training Programs. J Surg Educ. Aug 2023;80(8):1179–1187. doi:10.1016/j.jsurg.2023.05.015

18. Pretorius ES, Hrung J. Factors that affect National Resident Matching Program rankings of medical students applying for radiology residency. Acad Radiol. Jan 2002;9(1):75–81. doi:10.1016/s1076-6332(03)80298-2

19. Bernstein SA, Levy MS, McNeilly S, et al. Practice Location Preferences in Response to State Abortion Restrictions Among Physicians and Trainees on Social Media. J Gen Intern Med. Aug 2023;38(10):2419–2423. doi:10.1007/s11606-023-08096-5

20. Wright KM, Ryan ER, Gatta JL, Anderson L, Clements DS. Finding the Perfect Match: Factors That Influence Family Medicine Residency Selection. Fam Med. Apr 2016;48(4):279–85.

21. Galesic M, Bosnjak M. Effects of Questionnaire Length on Participation and Indicators of Response Quality in a Web Survey. Public Opinion Quarterly. 2009;73(2):349–360. doi:10.1093/poq/nfp031

22. Jebb AT, Ng V, Tay L. A Review of Key Likert Scale Development Advances: 1995-2019. Front Psychol. 2021;12:637547. doi:10.3389/fpsyg.2021.637547

23. Chryst B, Marlon J, Van Der Linden S, Leiserowitz A, Maibach E, Roser-Renouf C. Global warming’s “six Americas short survey”: Audience segmentation of climate change views using a four question instrument. Environmental Communication. 2018;12(8):1109–1122.

24. The jamovi project (2025). jamovi (Version 2.6) [Computer Software]. Retrieved from https://www.jamovi.org.

25. Lewandowski RE, Clayton SD, Olbrich L, et al. Climate emotions, thoughts, and plans among US adolescents and young adults: a cross-sectional descriptive survey and analysis by political party identification and self-reported exposure to severe weather events. The Lancet Planetary Health. 2024;8(11):e879–e893.

26. Heydari A, Partovi P, Zarezadeh Y, Yari A. Exploring medical students’ perceptions and understanding of the health impacts of climate change: a qualitative content analysis. BMC Med Educ. Oct 18 2023;23(1):774. doi:10.1186/s12909-023-04769-1

27. Leiserowitz A, Maibach E, Rosenthal S, et al. Global Warming’s Six Americas, Fall 2024. Yale University and George Mason University. New Haven, CT: Yale Program on Climate Change Communication.2025.

28. Carman J, Verner M, Rosenthal S, et al. Global Warming’s Six Audiences around the world. Accessed June 29, 2025. https://climatecommunication.yale.edu/publications/six-audiences-around-the-world/

29. Ayers M, Marlon JR, Ballew MT, et al. Changes in Global Warming’s Six Americas: an analysis of repeat respondents. Climatic change. 2024;177(6):96–96. doi:10.1007/s10584-024-03754-x

30. Motlogeloa O, Fitchett JM. Climate and human health: a review of publication trends in the International Journal of Biometeorology. Int J Biometeorol. Jun 2023;67(6):933–955. doi:10.1007/s00484-023-02466-8

31. Douris J, Kim G. The Atlas of Mortality and Economic Losses from Weather, Climate and Water Extremes (1970-2019). 2021;

32. Romanello M, Di Napoli C, Drummond P, et al. The 2022 report of the Lancet Countdown on health and climate change: health at the mercy of fossil fuels. The Lancet. 2022;400(10363):1619–1654.

33. Ccami-Bernal F, Barriga-Chambi F, Quispe-Vicuña C, et al. Health science students’ preparedness for climate change: a scoping review on knowledge, attitudes, and practices. BMC Med Educ. Jun 11 2024;24(1):648. doi:10.1186/s12909-024-05629-2

34. Khabour MO, Tarabsheh OO, Al-Zu’bi BM, Khabour OF, Saadeh R. Knowledge and attitudes of medical students towards the health impact of climate change: A study from Jordan. PLoS One. 2025;20(5):e0324943. doi:10.1371/journal.pone.0324943

35. Marill MC. Pressured By Students, Medical Schools Grapple With Climate Change. Health Aff (Millwood). Dec 2020;39(12):2050–2055. doi:10.1377/hlthaff.2020.01948

36. Omrani OE, Dafallah A, Paniello Castillo B, et al. Envisioning planetary health in every medical curriculum: An international medical student organization’s perspective. Med Teach. Oct 2020;42(10):1107–1111. doi:10.1080/0142159X.2020.1796949

37. Cerceo E, Marwah H, Philipsborn RP. Integrating Planetary Health Into Residencies: A Vital Step for Medical Education. Acad Med. Aug 08 2025;doi:10.1097/ACM.0000000000006184

38. Kircher M, Doheny BM, Raab K, Onello E, Gingerich S, Potter T. Understanding the knowledge, attitudes, and practices of healthcare professionals toward climate change and health in Minnesota. Challenges. 2022;13(2):57.

39. McCright AM, Dunlap RE. Cool dudes: The denial of climate change among conservative white males in the United States. Global environmental change. 2011;21(4):1163–1172.

40. Egan PJ, Mullin M. Climate change: US public opinion. Annual Review of Political Science. 2017;20(1):209–227.

