## Supplemental Table 1 for "Residency in the Era of Climate Change: A Multi-Institutional Survey of Medical Student Perceptions and Match Preferences"

**Table 1. Final Survey Questions:** Questions 1 - 2 assess student characteristics. Questions 3-6 assess the Six America’s group and are derived from SASSY. Questions 7-10 were newly developed for the present study to assess climate change and the impact on future career decisions.

| **Survey Question** | **Response Type** |
| --- | --- |
| 1. Year in medical school | 1, 2, 3, 4 |
| 2. What is your intended specialty? | Multiple choice (list of 25+ specialties + “Other”) |
| 3. How important is the issue of global warming to you personally? | Not at all – Extremely (5-point Likert) |
| 4. How worried are you about global warming? | Not at all – Very (4-point Likert) |
| 5. How much do you think global warming will harm you personally? | Not at all – A great deal (5-point Likert) |
| 6. How much do you think global warming will harm future generations of people? | Not at all – A great deal (5-point Likert) |
| 7. Climate change is an important threat to human health. | Strongly Disagree – Strongly Agree (5-point Likert) |
| 8. In choosing my medical specialty, I consider the relative importance of climate change and the potential impact it will have on my career and patients. | Strongly Disagree – Strongly Agree (5-point Likert) |
| 9. It is important to me that my residency program acknowledges the health impact of climate change and demonstrates commitment to sustainability. | Strongly Disagree – Strongly Agree (5-point Likert) |
| 10. The level of dedication to sustainability and climate change mitigation will impact my preference list of residency programs. | Strongly Disagree – Strongly Agree (5-point Likert) |
